# Intravenous Antibiotics in Preterm Infants have a Long-Term Negative Effect upon Microbiome Development Throughout Preterm Life – an observational study

**DOI:** 10.1101/2022.09.29.22280493

**Authors:** RA Hutchinson, KL Costeloe, WG Wade, MR Millar, K Ansbro, F Stacey, PF Fleming

## Abstract

Intestinal dysbiosis is implicated in the origins of necrotising enterocolitis and late-onset sepsis in preterm babies. However, the effect of modulators of bacterial growth (*e*.*g*. antibiotics) upon the developing microbiome is not well-characterised.

Using high-throughput 16S rRNA gene sequencing combined with contemporaneous clinical data collection, the within-subject relationship between antibiotic administration and microbiome development was assessed, in comparison to preterm infants with minimal antibiotic exposure.

During courses of antibiotics, diversity progression fell in comparison to that seen outside periods of antibiotic use (−0.71units/week *vs*. +0.63units/week, p<0.01); *Enterobacteriaceae* relative abundance progression conversely rose (+10.6%/week *vs*. -8.9%/week, p<0.01). After antibiotic cessation, diversity progression remained suppressed (+0.2units/week, p=0.02).

Antibiotic use has an acute and longer-lasting impact on the developing preterm intestinal microbiome. This has clinical implications with regard to the contribution of antibiotic use to evolving dysbiosis, and affects the interpretation of existing microbiome studies where this effect modulator is rarely accounted for.

## Introduction

Intestinal dysbiosis is increasingly implicated in the pathophysiological origins of preterm neonatal conditions such as necrotising enterocolitis (NEC) and late-onset sepsis (LOS), leading to an interest in understanding the development of the microbiome in these settings.Technologies such as high-throughput 16S rRNA gene sequencing have allowed an enhanced depth and breadth of microbial analyses^1-3^. Considering the evolution and variation in neonatal practice across time and between units, it is unsurprising that variable patterns of microbiome development have been described across studies^4,5^. Some consistent trends have however been identified and replicated using different methodologies. Typically, that after an initial period of dominance by staphylococci, the relative abundance of *Enterobacteriaceae* increases, subsequently supplanted by a variety of (often anaerobic) taxa into later preterm life^1,3,6,7^. Overall diversity of the bacterial community increases with postnatal age.

The effect of potential modifiers of microbiome development encountered during the course of preterm neonatal care is, however, not well understood, nor accounted for in analyses; antibiotics represent one such intervention. Numerous studies have demonstrated differences in microbiome development in antibiotic-exposed infants, such as a reduction in the relative abundance of typical commensal organisms (*e*.*g*. bifidobacteria, lactobacilli)^8-12^; and an increased relative abundance of taxa within the Proteobacteria phylum, particularly the family *Enterobacteriaceae*^1,2,13,14^. It is not possible, however, to delineate a temporal relationship between antibiotic use and associated dysbiosis.

A barrier to understanding this association lies in the manner in which analyses occur at population-level, rather than on an individual subject level, where the relationship between the stimulus (antibiotic use) and response (dysbiosis) can be more readily elucidated. Amalgamation of analyses from these populations of infants with their potentially diverse microbial communities^11,12^, and without accounting and controlling for these potential confounders may lead to misleading conclusions. If the associations described in population-level analyses *are* legitimately causative, this is of important clinical relevance, in light of the widespread use of antibiotics in the preterm population^15^. The aim of this study was to evaluate these purported associations through a high-fidelity assessment of intestinal microbiome development in preterm infants on an individual subject-level, during predefined episodes of purported clinical and microbiome stability, and discrete episodes of antibiotic administration.

The shift towards increased proportions of potentially pathogenic *Enterobacteriaceae* and reduced commensal organisms is at the heart of the dysbiotic origins of the proposed mechanisms of NEC and LOS in preterm infants. Commensal organisms augment gut health through enhanced intestinal activity and growth^16,17^; improved luminal barrier integrity^16^; and immune-tolerance induction^18,19^; with their reduction, these actions are attenuated. Conversely, *increased* proportions of *Enterobacteriaceae* heighten the risk of interaction between elevated TLR4 levels within the preterm gut^20^ and Gram-negative, lipopolysaccharide-expressing bacteria, resulting in downregulation of intestinal growth^21^, ischaemia^22^ and ultimately catastrophic immune activation.

## Results

### Study population and samples

There were 158 eligible infants admitted to the neonatal unit during the study period; of these 76 were recruited into the study after informed consent. Five subjects did not continue in the study long enough to have a stool sample collected (due to withdrawal, transfer out of the unit, or death), so microbiome composition data from 71 subjects were analysed (see Fig.1). Of these 71 infants, 35 met the eligibility criteria for inclusion in Groups 1 or 2a/b; the remaining 36 infants are not discussed in this paper.

**Fig. 1.**
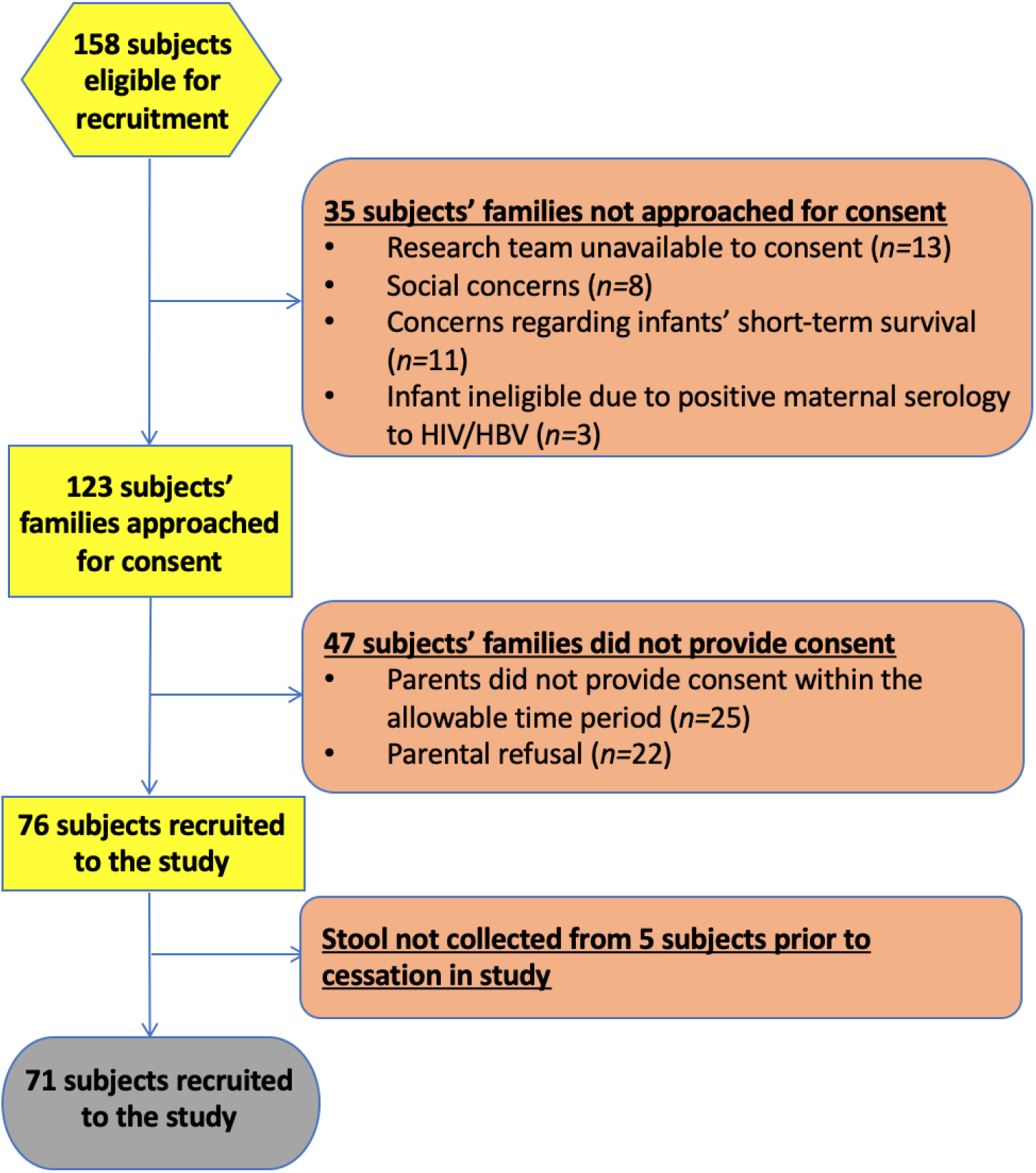
Recruitment Flowchart

14 subjects were assigned to Group 1 (‘minimal antibiotic-exposure’), with one subsequently being retrospectively excluded on the basis of their incongruence with the remainder of the group (*i*.*e*. uniquely a positive history of chorioamnionitis; significantly lower birthweight and gestational age; and early study cessation). 21 subjects were eligible for Group 2a (‘during antibiotics’); of those, 16 remained eligible to be included in Group 2b (‘post-antibiotics’). Subject demographics are outlined in Table 1.

**Table 1.** Demographics of Subjects.

|  | Group 1<br>Total (n=13) | Excluded<br>subject | Group 2a<br>Total (n=21) | Group 2b<br>Total (n=16) |
| --- | --- | --- | --- | --- |
| **Gestational age**, mean (SD), *range*, weeks | 30 (0.8), 29.1-31.7 | **26.4** | 27.7 (2.3), 23.6-31.3 | 28.3 (2.2), 24.9-31.3 |
| **Birthweight**, mean (SD), *range*, grams | 1347 (215), 1020-1780 | **1050** | 957 (294), 600-1835 | 982 (316), 600-1835 |
| **Gender**, n'/n | 7/13 (54%) Male | *Female* | 12/21 (57%) Male | 9/16 (56%) Male |
| **Plurality of pregnancy**, n'/n | 5/13 (38%) singletons<br>7/13 (54%) twins (inc. two pairs)<br>1/13 (8%) triplet | *Singleton* | 17/21 (81%) Singletons<br>2/21 (10%) Twins<br>2/21 (10%) Triplets | 13/16 (81%) Singletons<br>2/16 (13%) Triplets<br>1/16 (6%) Twins |
| **History of abnormal Dopplers**, n'/n | 5/13 (38%) | *No* | 3/21 (14%) | 3/16 (19%) |
| **Small for gestational age** n'/n | 1/13 (8%) | *No* | 6/21 (29%) | 6/16 (38%) |
| **Prolonged rupture of membranes prior to delivery**, n'/n | 2/13 (15%) | **Yes** | 7/21 (33%) | 4/16 (25%) |
| **Antenatal steroids completed >24 hours prior to delivery**, n'/n | 9/13 (69%) | *Yes* | 14/21 (67%) | 11/16 (69%) |
| **Associated diagnosis of chorioamnionitis**, n'/n | 0/13 (0%) | **Yes** | 8/21 (38%) | 5/16 (31%) |
| **Maternal peripartum antibiotic use**, n'/n | 4/13 (31%) | *Yes* | 4/21 (19%) | 4/16 (25%) |
| **Location of delivery**, n'/n | 13/13 (100%) Inborn | *Inborn* | 20/21 (95%) inborn | 16/16 (100%) inborn |
| **Primary cause of preterm birth**, n'/n | 5/13 (38%) Pregnancy-induced hypertension<br>4/13 (31%) Poor fetal growth<br>2/13 (15%) PPROM<br>1/13 (8%) APH<br>1/13 (8%) Poor fetal growth in sibling | *PPROM* | 6/21 (29%) PPROM<br>5/21 (24%) Preterm labour without PPROM<br>4/21 (19%) Pregnancy-induced hypertension<br>2/21 (10%) Poor fetal growth<br>2/21 (10%) Poor fetal growth in sibling<br>1/21 (5%) APH<br>1/21 (5%) Decreased fetal movements | 4/16 (25%) Preterm labour without PPROM<br>4/16 (25%) Pregnancy-induced hypertension<br>3/16 (19%) PPROM<br>2/16 (13%) Poor fetal growth<br>1/16 (6%) Poor fetal growth in sibling<br>1/16 (6%) APH<br>1/16 (6%) Decreased fetal movements |
| **Apgar score at 5 mins**, median, *range* | 9, 6-10 | *6* | 9, 2-10 | 9, 2-10 |
| **Worst BE within first hour**, median, *range* | -4.3, -11.2 – -2.2 | *-3.5* | -4.7, -17.3 – -1 | -4.6, -9 – -1 |
| **Temperature on admission**, mean, *range*, °C | 36.5, 35-38.3 | *37.1* | 36.8, 36-37.8 | 36.8, 36-37.8 |
| **Mode of delivery**, n'/n | 13/13 (100%) Operative | **Vaginal** | 13/21 (62%) Operative | 11/16 (69%) Operative |

Subjects in Group 1 received antibiotics for a median of four days. The courses of antibiotics in the subjects in Group 2a commenced at a mean corrected gestational age of 31^+2^/40 and lasted for a median of seven days. The period post-antibiotics in Group 2b commenced at a mean corrected gestational age of 32^+1^/40.

2229 samples were obtained from subjects during the course of the study (median 30 samples/subject). In view of restrictions on numbers of samples, 1434 progressed to DNA extraction. Following exclusion of samples without detectable DNA following PCR and removal of samples with <5000 sequences, 1193 samples remained (19 samples/subject). Of the 14 subjects eligible for inclusion in Group 1, after exclusion of the one clinical outlier (described above), analyses were conducted on 233 samples (median 19/subject). Of the 21 subjects with clinical episodes eligible for inclusion in Group 2a, analyses were conducted on 93 samples (median 4/subject). Of the 16 subjects with clinical episodes eligible for inclusion in Group 2b, analyses were conducted on 133 samples (median 8/subject).

### Microbiome Progression in Group 1 (minimal antibiotic exposure) – see Fig.2

Diversity was seen to increase proportionally with postnatal age, with individual trajectories ranging from 0.15-1.27 units/week. The weighted mean rate of diversity progression in Group 1 was 0.63units/week (weighted 95%CI=0.44-0.82).

The most abundant family-level taxon in Group 1 was *Enterobacteriaceae* (with a weighted, normalised median AUC of 70.3% [95%CI = 58.4 – 78.5%]) which was seen ubiquitously across all subjects and samples. *Enterobacteriaceae* was seen to fall from a peak abundance (weighted median 96.3% [95.3-99.6%] at around an average of Day 13 [10-16]), at a mean rate of decline of -8.9%/week (95%CI = -5.7 – -12.2) – see Fig.2.

**Fig.2.**
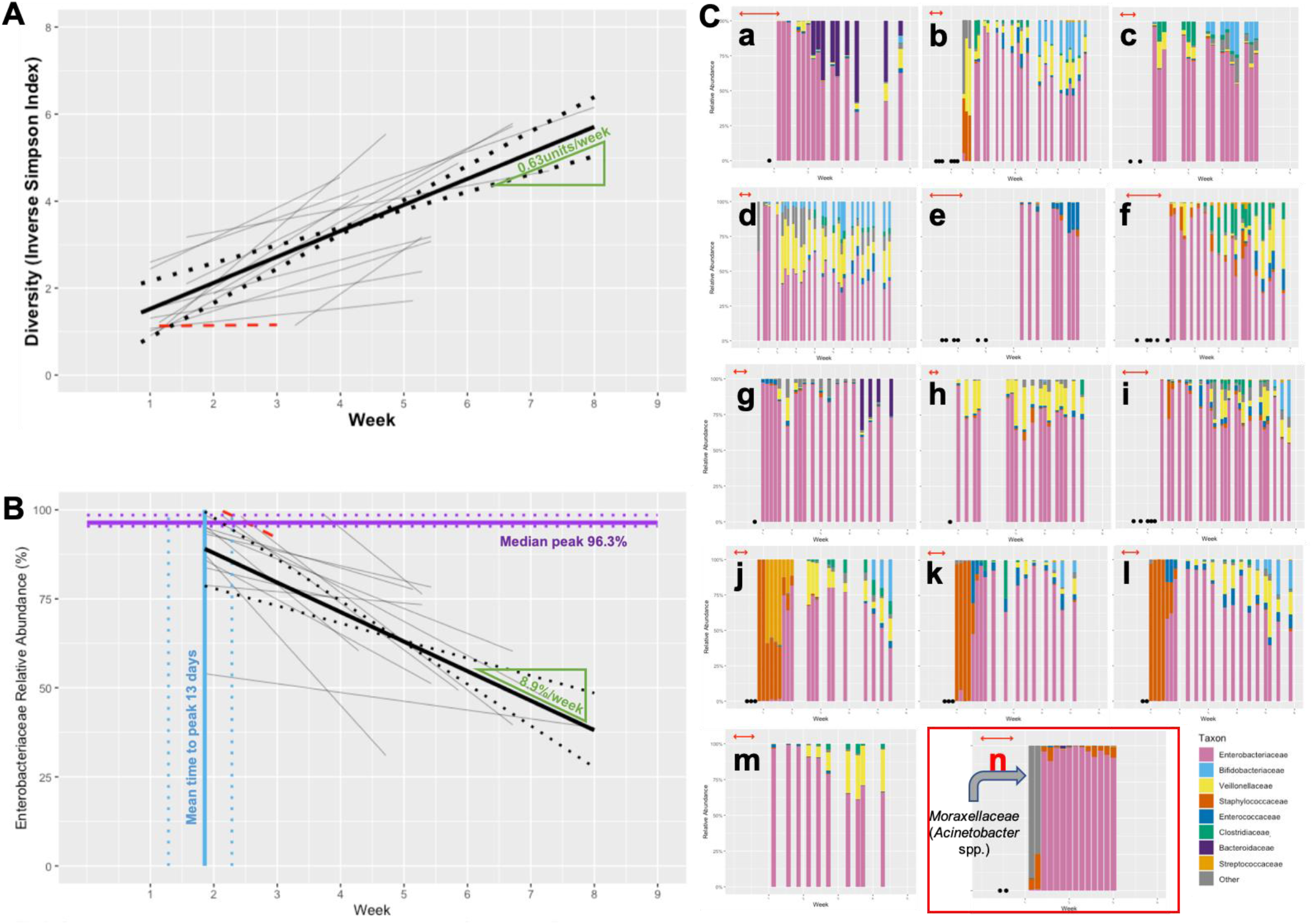
**Fig.2A**Composite plot of individual trajectories of longitudinal diversity progression in stable babies minimally-exposed to antibiotics (Group 1) shown in grey. The subject with chorioamnionitis who was excluded from all calculations of mean trajectories is shown in red. Weighted mean trajectory (thick black line) – gradient labelled in green. 95%CI for weighted mean trajectory denoted by dotted lines **Fig.2B** Composite graph demonstrating the three components of assessed *Enterobacteriaceae* relative abundance progression in stable infants minimally-exposed to antibiotics (Group 1): weighted median peak relative abundance (purple); weighted mean time to peak relative abundance (blue); weighted mean regression coefficient of fall from peak (black, with 95%CI denoted by accompanying dotted lines), with gradient shown in green; individual weighted regression coefficients from which the mean is derived (grey). As in Fig. 2A, data from the excluded infant with chorioamnionitis (red) is not included in the calculations. **Fig.2C** Longitudinal taxonomic profiles of the majority taxa in stable babies minimally-exposed to antibiotics (Group 1; a-m), and the excluded subject (n), highlighted in red. Horizontal red arrows indicate periods of antibiotic administration; black spots on horizontal access represent samples from which no DNA was extracted.

In contrast with the falling relative abundance over time of *Enterobacteriaceae*, other taxa were seen to develop within the intestinal bacterial community in Group 1 as postnatal age increased. *Veillonellaceae* (10.4% [4.5-18.0%]) and *Enterococcaceae* (3.3% [0.9-4.3%]) were seen in samples across all subjects in the group, and increased in relative abundance proportionally with postnatal age (*Veillonellaceae* weighted median increase of 1.4%/week [95%CI 0.01-3.7]; *Enterococcaceae* 0.4%/week [0.07-1.3]).

Other taxa were also seen in significant median relative abundances (*Staphylococcaceae* 2.9%; *Bifidobacteriaceae* 2.5%; *Clostridiaceae* 2.3%), but were either inconsistently present across subjects within the group, or displayed no trends, thus precluding group-level quantitative descriptions. However, despite their relative low median abundances, at times these taxa represented significant proportions of the intestinal bacterial community. *Staphylococcaceae* were temporally skewed to samples from an early postnatal age, where they could represent nearly 100% of the detected taxa; *Bifidobacteriaceae* were conversely skewed to the later preterm period, but could represent >25% of detected taxa at these times. Indeed, some taxa (i.e. *Streptococcaceae* and *Bacteroidaceae*) with a very low median abundance across the cohort (<1%, and not further analysed), comprised, in two individuals, a significant proportion of the microbiome (*i*.*e*. 9% and 28%, respectively). Features such as these show that alongside some consistent trends in preterm intestinal microbiome development (*i*.*e*. diversity progression, and *Enterobacteriaceae* decline), there remain some periods of profound dynamic change in taxa, and significant inter-subject variability – see Fig.2.

### Microbiome Progression in Group 2a (during antibiotics)

Standard first-line antibiotics for late-onset sepsis at Homerton University Hospital NICU were flucloxacillin and gentamicin, with ceftazidime and vancomycin as a second-line. Flucloxacillin and penicillin + gentamicin combinations have been associated with increased relative abundance of *Enterobacteriaceae* in the intestinal microbiome^28,29^. Conversely, ceftazidime is associated with reduced *Enterobacteriaceae*^*30*^ and vancomycin with reduced *Staphylococcaceae*^*28*^.

During a course of antibiotics, diversity was seen to *fall* by -0.71units/week (95%CI = -0.24 – -1.2). When compared against the baseline rate progression established in Group 1 (a *rise* 0.63units/week), where there was minimal antibiotic exposure, this was shown to differ significantly (*p*«0.001). Similarly, the decline in *Enterobacteriaceae* relative abundance seen in Group 1 (−8.9%/week) was reversed during a course of antibiotics, and increased by 10.6%/week (95%CI = 1.1-30.2), which was again noted to differ significantly (*p*«0.001). These findings demonstrate the acute effect of antibiotic administration on the gut flora – see Fig.3.

**Fig.3.**
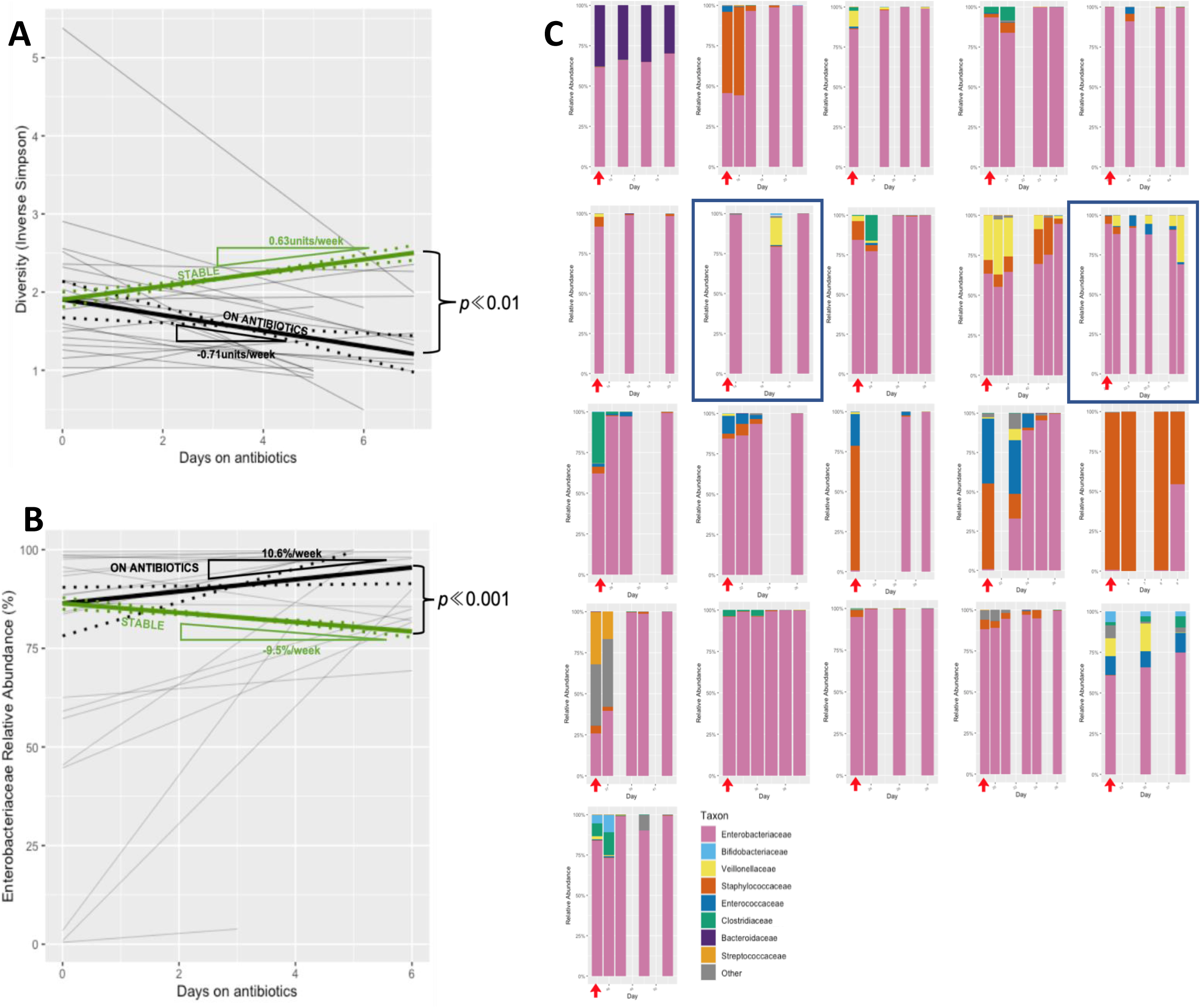
**Fig.3A**Composite plot of individual trajectories of longitudinal diversity progression during antibiotic administration (Group 2a) shown in thin grey lines, with weighted mean trajectory (thick black line). For comparative purposes, trajectory of diversity progression in Group 1 overlaid (thick green line). 95%CI for weighted mean trajectories denoted by dotted lines. For clarity and comparative purposes, the intercept of the two trajectories is plotted at the median intercept of the group on antibiotics **Fig.3B** Composite plot of individual trajectories of longitudinal *Enterobacteriaceae* relative abundance progression during antibiotic administration (Group 2a) shown in thin grey lines, with weighted median trajectory (thick black line). For comparative purposes, median trajectory of *Enterobacteriaceae* relative abundance progression in Group 1 overlaid (thick green line). 95%CI for weighted median trajectories denoted by dotted lines. For clarity and comparative purposes, the intercept of the two trajectories is plotted at the median intercept of the group on antibiotics. **Fig.3C** Longitudinal taxonomic profiles of the majority taxa in babies during periods of receiving intravenous antibiotics (Group 2a). Red arrows indicate the sample taken prior to commencing antibiotics. Antibiotic courses containing ceftazidime highlighted in blue.

**Fig.4.**
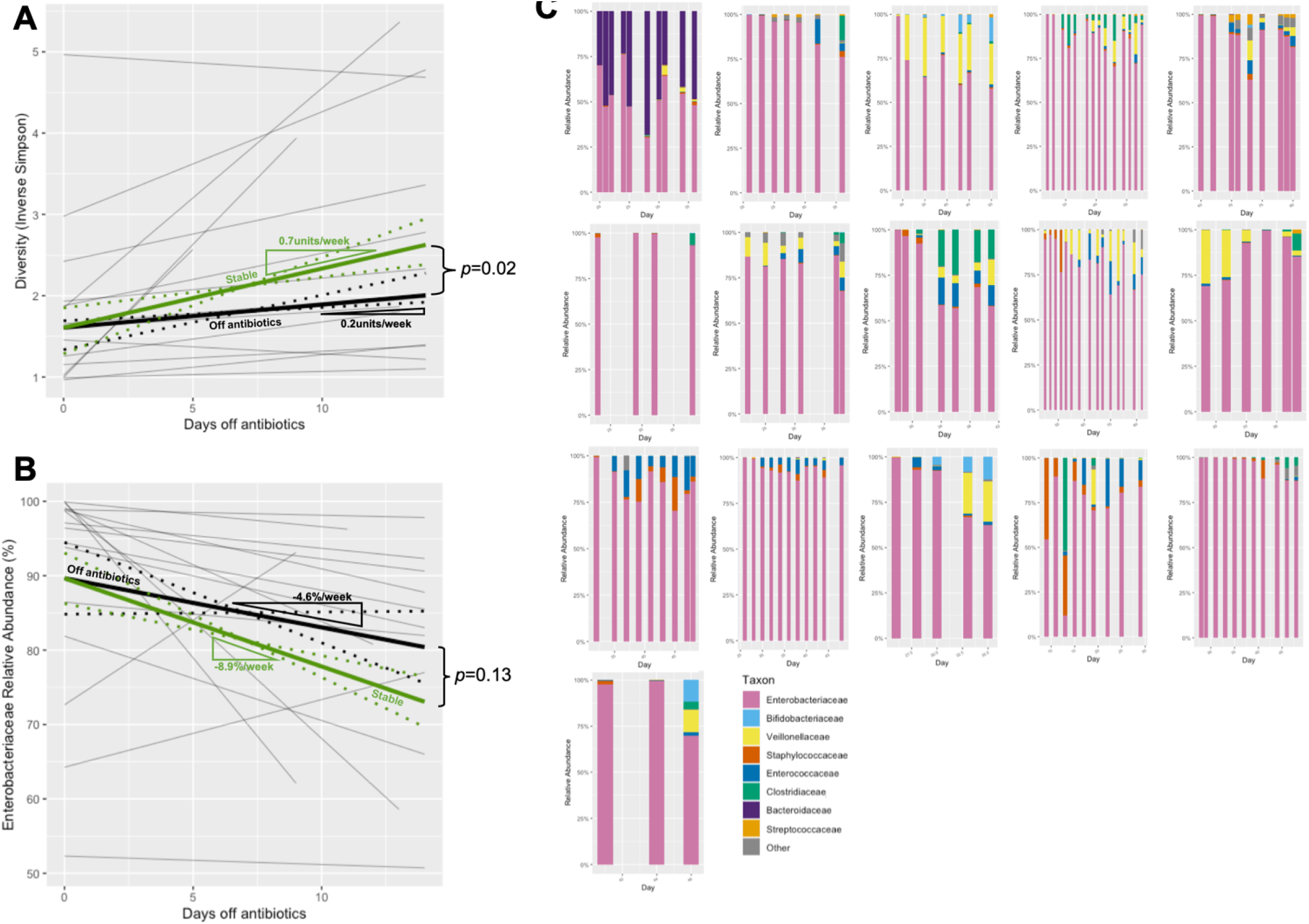
**Fig.4A**Composite plot of individual trajectories of longitudinal diversity progression following antibiotic cessation (Group 2b) shown in thin grey lines, with weighted median trajectory (thick black line). For comparative purposes, trajectory of diversity progression in stable subjects (Group 1) overlaid (thick green line). 95%CI for weighted median trajectories denoted by dotted lines. For clarity and comparative purposes, the intercept of the two trajectories are both plotted at the median *y*-intercept of Group 2b. **Fig.4B** Composite plot of individual trajectories of *Enterobacteriaceae* relative abundance progression following antibiotic cessation (Group 2b) shown in thin grey lines, with weighted mean trajectory (thick black line). For comparative purposes, trajectory of *Enterobacteriaceae* relative abundance progression in stable subjects (Group 1) overlaid (thick green line). 95%CI for weighted mean trajectories denoted by dotted lines. For clarity and comparative purposes, the intercept of the two trajectories are both plotted at the median *y*-intercept of Group 2b. **Fig.4C** Longitudinal taxonomic profiles of the majority taxa in babies during periods after cessation of intravenous antibiotics.

In view of the contrasting hypothesised effects of the differing antibiotics combinations used, a *post-hoc* analysis was undertaken examining the taxonomic progression during courses of antibiotics where ceftazidime was employed. Whilst only small numbers, as predicted by previous studies the *Enterobacteriaceae* relative abundance *fell* by -15.9%/week, in contrast to the pattern seen when gentamicin was used as the component providing Gram-negative cover.

As anticipated by the noted falling diversity and rising *Enterobacteriaceae* relative abundance, a mixture of other taxa (reflective of the inter-subject variability demonstrated in Group 1) were largely eradicated to leave a heavily *Enterobacteriaceae*-dominated bacterial community. There were some notable individual exceptions where a more diverse microbiome was maintained despite the antibiotics – whether this was reflective of the resilience of the microbiome, or represented a host factor, is not known.

### Microbiome Progression in Group 2b (post-antibiotics)

Following the cessation of antibiotics, diversity progression resumed at 0.2units/week (95%CI = 0.11-0.47); although, in comparison to the progression in Group 1 subjects, this was diminished (0.2 *vs*. 0.7units/week [*n*.*b*. median values], *p*=0.02). Similarly, the *fall* in *Enterobacteriaceae* relative abundance resumed following the cessation of antibiotics (--4.6%/week [0.4 – -9.6%]). Whilst this was reduced in comparison to the rate seen in Group 1 subjects (−4.6 *vs*. -8.9%/week), this was not statistically significant (*p*=0.13).

The relative abundance of other taxa was also examined in the post-antibiotic period. In light of the preponderance of *Enterobacteriaceae* resulting from the preceding course of antibiotics, and compounded by the shallower rate of decline in Group 2b, other taxa were correspondingly reduced compared to Group 1. Of the main taxa analysed, *Veillonellaceae* (10.4% [in Group 1] *vs*. 2.4% [in Group 2]), *p*=0.02; *Clostridiaceae* (2.3% *vs* 0.39%, *p*=0.04) and *Bifidobacteriaceae* (2.5% *vs*. 0.006%, *p*<0.0001) were seen to be significantly reduced in the post-antibiotic period.

## Discussion

That antibiotics impact upon preterm intestinal microbiome development is predictable; supported by clinical correlates from adult medicine and is seen in a number of studies in preterm infants. However, this paper quantitatively describes this relationship on an individual subject level, using strict criteria to analyse this effect in isolation from other clinical factors.

We have demonstrated that during a course of intravenous antibiotics (Group 2a), diversity fell at a rate of 0.71units/week, a significant difference from the rise seen in infants only minimally-exposed to antibiotics (Group 1) and in keeping with observations from the published literature^14,31,32^. Notably, the development of NEC has been shown to be associated with both reduced microbial diversity *and* prior antibiotic use^33,34^; our finding may explain these associations. It may be that the reported association between NEC and prior antibiotic use merely reflects a confounding underlying pathophysiological process which represents a precursor to NEC, *and* which also induces clinical features which may prompt antibiotic use. However, it may be that there is a true causative mechanism, whereby the use of antibiotics reduces the diversity of bacteria present, with consequent reduction of the benefits to intestinal health which are conferred from the normal array of commensal organisms.

It was also noted that *Enterobacteriaceae* relative abundance rose at a rate of 10.6 %/week whilst receiving antibiotics – a significant change from the progression in antibiotic-naïve subjects; this is a feature that has been described elsewhere in the literature^1,6,14^. It is important to note that this does not imply that *Enterobacteriaceae* growth was augmented – studies have demonstrated a general *reduction* in absolute total bacterial abundance under the action of antibiotics^35^. Rather, this implies that whilst *Enterobacteriaceae* reduced in total abundance under the action of antibiotics, this occurred at a lower rate than for other taxa, leading to an increase in *relative* abundance of *Enterobacteriaceae*. An increased relative abundance of γ -Proteobacteria (the class to which *Enterobacteriaceae* belong) has been associated with an increased risk of the development of NEC^36,37^. It may be hypothesised that increased proportions of endotoxin-expressing Gram-negative organisms (*e*.*g. Enterobacteriaceae*) within the intestinal lumen increase the risk of inappropriate stimulation of TLR4-replete immune cells within the gut^38^, precipitating an inflammatory cascade culminating in clinical NEC. However, the commensal nature of many commonly-encountered Gram-negative organisms encountered within the gut implies that this may only represent one component of the development of a pathogenic interaction.

Uniquely, the development of the microbiome *after* a course of antibiotics was also examined in this study. It was seen that following the cessation of antibiotics, the accrual of increasing diversity was again seen, although at a significantly reduced rate compared to antibiotic-naïve subjects (0.2 *vs*. 0.7 units/week, *p*=0.01). Similarly, the fall in relative abundance of *Enterobacteriaceae* recurred, again at a reduced rate compared to the antibiotic-naïve cohort (−4.6 *vs*. -8.6%/week - although statistical significance was not met [*p*=0.13]). The reasons for this may be considered to result from both components of the microbiome: the inhabitants and the environment. In terms of the inhabitants, it may reflect that microbiome development is driven, at least in part, by its founder organisms. In this instance, the residual post-antibiotic microbiome is heavily *Enterobacteriaceae*-dominated; from this state, once the negative modulator of development (*i*.*e*. antibiotics) is removed, the microbiome regrows, but retains its *Enterobacteriaceae*-dominance, due to a paucity of alternative founder organisms and loss of an ecological niche for them. Additionally, the intestinal environment within which the bacteria reside may have been altered by the preceding change in bacteria, or the clinical episode which precipitated the antibiotic use. Commensal organisms have a self-sustaining effect within the intestinal lumen (*i*.*e*. induction of commensal-specific nutrient expression by enterocytes and commensal-sparing anti-microbial production^39,40^) – their eradication means these mutualistic mechanisms may be lost, leading to the gut becoming less hospitable to these organisms, with persistence of residual *Enterobacteriaceae* at higher proportions. Similarly, if the preceding antibiotic course had been mandated by an episode of sepsis, the immune-profile of the infant and their intestine may have been altered by this, to a more pro-inflammatory state in which the organisms which could have been hoped to supplant the dominance of *Enterobacteriaceae* cannot flourish.

The effect of antibiotics upon other taxa was also examined. In conjunction with *Enterobacteriaceae*-dominance and augmented persistence even after stopping antibiotics, there was a corresponding reduction in the non-*Enterobacteriaceae* organisms normally seen to inhabit the preterm gut. Whilst their uneven distribution across subjects precluded longitudinal analyses in the manner possible for *Enterobacteriaceae*, gross differences could still be seen. The most significant differences were seen in the relative abundance of *Veillonellaceae, Clostridiaceae* and *Bifidobacteriaceae*, the latter of which showed a 400-fold fall in relative abundance compared to that seen in infants minimally-exposed to antibiotics. It has been previously demonstrated that bifidobacteria levels fall in some subjects receiving antibiotics^9,10^, but that this persists even after antibiotics have ceased, is a novel finding. It may be hypothesised that bifidobacteria are especially sensitive to the aforementioned mechanisms of commensal suppression, either through particular sensitivity to the antibiotic combinations employed in this neonatal population, or through the intestinal environment changing in a manner which makes it inhospitable to bifidobacterial colonisation. This finding is of importance, considering the frequent proposition of the role of bifidobacteria in preterm probiotic therapies. Their acute sensitivity to commonly-used antibiotics and sustained reduction even after the cessation of antibiotics may explain their variable success in clinical trials^41^, with successful colonisation seemingly heavily influenced by underlying antimicrobial practice.

At the time of conception and undertaking of this study, 16S rRNA gene sequencing represented the methodology of choice to study the components of diverse microbiota. However, the advent of shotgun metagenomics, where the *entire* target genome is analysed (in comparison to just the 16S rRNA gene), now allows greater depth of taxonomic differentiation. Within this study, taxa were analysed at family-level, as this represented the taxonomic level which maximised depth, without excluding large numbers of unclassified sequences (*i*.*e*. 99.5% of sequences were classified to family-level; only 73.8% were classified to genus-level) – this was largely due to homogeneity within the 16S rRNA gene V4 hypervariable region amongst *Enterobacteriaceae*. Whilst this was pragmatically necessitated, this depth of differentiation did not reach the level of maximum clinical applicability, when antibiotic choices are based upon species and resistance profiles, and probiotics are formulated at strain level^42^. The capability of shotgun metagenomics to differentiate to these levels has been demonstrated^43-45^ and is of the utmost relevance when a clinical future of microbiome manipulation can be envisaged, via the use of narrow-spectrum, targeted antibiotic regimens and probiotic administration.

Whilst the sub-group of infants minimally-exposed to antibiotics (Group 1) has been pragmatically used as a ‘control’ group for comparative purposes in this study, this does not account for other modulators of microbiome progression. Factors which may reasonably be hypothesised to affect microbiome development include the maternal/parental microbiome (both in terms of founder organisms, and subsequent exposures) and diet. It is also possible that there exist numerous influences which cannot be anticipated. True matching for all potential modulators is likely prohibitively restrictive; a solution may lie in larger scale studies (in terms of numbers of subjects) where these confounders may be more easily balanced, or in paired analyses around an intervention, where the subject acts as their own control.

This study has demonstrated that the impact of antibiotics upon the developing preterm intestinal microbiome is seen not just during the acute course of antibiotics, but remains throughout preterm life. In light of the association between this dysbiosis and the development of important neonatal conditions such as NEC and LOS, this finding may provide further support for the judicious use of antibiotics in this vulnerable patient group, or more narrow-spectrum regimes which do not have such a dysbiotic effect.

Additionally, we have demonstrated the value and feasibility of this novel application of an individual subject analysis to microbiome assessment. The assessment of babies, based purely upon a clinical categorisation (*e*.*g*. prematurity) without accounting for the effects that underlying factors (*e*.*g*. antibiotics use) may have, potentially leads to a skewed interpretation of a population’s apparent microbiome development. This methodology may provide a blueprint for further studies to account for such confounding factors in future work.

## Methodology

### Ethical Approval, Funding and Recruitment

This study was nested within the wider observational study “Investigating Microbial Colonisation and Immune Conditioning in Preterm Babies”^23^ and was prospectively incorporated into its design and regulatory approvals. Preterm infants were recruited at Homerton University Hospital (London, UK), between 23^+0^–31^+6^/40 weeks, within the first 72hrs after birth, between January 2016 and March 2017. Ethical approval was granted by the London (Chelsea) Research Ethics Committee (Ref: 15/LO/1924). The study was funded with a grant from Barts Charity (Ref: 764.2306).

### Sample Collection, Storage and Transfer

Stool samples were prospectively collected from birth up to the cessation of the subjects’ participation in the study (37/40 weeks CGA, or 12 weeks of age, whichever was sooner). Samples (when available) were taken on a daily basis within four hours of passage and immediately placed in a universal storage container (without preservatives) in a refrigerator at 4°C. The samples were subsequently transferred to an ultra-low freezer (−80°C) generally within 24 hours. Samples were later transferred from the clinical site to the laboratory for long-term storage and subsequent analyses; this was performed at intervals using a specialist medical courier service, with maintenance of the cold-chain throughout achieved through transport on dry ice (at -79° C). Duration of time in -80° C storage ranged from 5-20 months.

### Sample Selection

2229 samples were collected in total from all 71 subjects in this study; this number of samples exceeded the number of slots available for sequencing. Constraints on time for extraction; indexing capacity (384 [24×16] combinations from 40 [16+24] indexed primers, across four 96 [8×12] well PCR plates); and the financial cost of sequencing runs (four runs costed for) meant that 1536 samples could be sent for sequencing. As a quality control measure, slots were reserved for positive controls, negative controls and mock communities. Additionally, for other investigations not pertaining to this study, slots were reserved for PMA-untreated samples and *Salinibacter ruber* spiked samples. Consequently, 1434 samples were selected for analyses.

Samples were prospectively selected from those available on a pragmatic basis, aiming for:

- regularity of sampling (every 2-3 days, where able and available)
- the capture of episodes of potential microbiome instability in greater detail – up to daily sampling (*i*.*e*. early life, before/during/after courses of antibiotics, feeding/respiratory support changes, blood transfusions)
- the capture of ‘stable’ infants’ microbiome profiles in greater detail (in order to develop a reliable baseline

Where samples were unavailable/unsuitable for analysis (*e*.*g*. small volume samples; samples depleted in pilot studies), an alternative (temporally adjacent) sample was selected.The microbiome composition of samples was not known prior to selection.

### Library Preparation

The DNA library was prepared as per the recommended protocol for the DNeasy® PowerSoil® Kit (Qiagen, Netherlands), with the following adaptations:-

- a reduction in starting stool mass from 250mg to 20mg, following evidence from the literature, corroborated by our own pilot study, demonstrating improved extraction yields at reduced volumes^46^
- the addition of a manual homogenisation step, in view of viscous samples
- the use of propidium monoazide (PMA) following the standard protocol (Biotium, USA). PMA is a photo-reactive, DNA-binding dye, used to prevent downstream amplification of free DNA (*e*.*g*. contaminant, non-viable) in PCR^47,48^.

The 16S rRNA gene V4 region was amplified using Fusion primers (v4.SA501-508, v4.SB501-508, v4.SA701-712, v4.SB701-71; Eurofins, Germany) – the V4 region was utilised as it was theorised to offer the greatest capacity for differentiation between taxa, in view of it being one of the longest hypervariable regions. Samples from which no amplified DNA was subsequently demonstrated (using electrophoresis) were removed from further analyses. Normalisation of amplified DNA was undertaken as per the standard protocol for the SequalPrep™ Normalization Plate (96) Kit (Invitrogen, USA). DNA quantitation was carried out as per the standard protocol for Quant-iT™ PicoGreen® dsDNA Reagent (Invitrogen, USA).

### Sequencing and Processing

Amplicon sequencing was performed by the Barts and the London Genome Centre, using Illumina MiSeq Technology (2×250bp flow-cell for paired-end sequencing with 5% PhiX DNA). Processing of the sequencing data was conducted within R^49^ using the DADA2 pipeline^50^, using the standard protocol (*i*.*e*. truncations at position 240 for forward reads, 160 for reverse reads; truncation at a base quality score of 2; no ambiguous bases; maximum 2 expected errors; and automated removal of PhiX DNA). Batch processing was employed, based upon the originating sequencing run, due to the potential for run-specific errors and biases. Positive mock community controls were used to ensure sequencing accuracy (ZymoBIOMICS, USA). Amplicon sequence variants (ASVs) identified by the DADA2 algorithm were taxonomically-assigned using a bespoke, curated, site-specific (*i*.*e*. preterm gut) database of 16S rRNA gene sequences^51^. The *decontam* package^52^ in R was used to identify contaminant sequences (sequences disproportionately represented in negative controls); these sequences were manually reviewed and discarded if microbiologically-plausible as a contaminant.

Samples with <5000 sequences were discarded, in view of a risk of under-representing true richness and diversity.

### Statistics

Diversity was measured using the inverse Simpson index^25^. The central tendency of taxa varying in relative abundance over time were described using the area-under-the curve (AUC), normalised for duration in study^26^. Analyses and descriptive statistics were weighted, where possible, by number of reads/sample and number of samples/subject. Siblings from a multiple (twin) pregnancy were half-weighted (if within the same sub-group), in anticipation of non-independence in microbiome development. In light of the potential for multiple comparison errors from assessment of many taxa, analyses occurred at a family-level (this additionally allowed the inclusion of the greatest proportion of amplicon sequence variants [ASVs] in analyses); and only taxa with a median, normalised AUC >1% relative abundance were analysed. In anticipation of the longitudinal nature of analyses, where approximately linear trends were identified, individuals’ summary data were represented by the Theil-Sen regression coefficient^27^. Without a precedent of similar longitudinal microbiome studies, a pragmatic decision was made to aim for an 80% power to detect a one standard deviation difference in diversity progression between groups; this would require 16 subjects/group. It was prospectively noted that subject numbers would be restricted by an absence of an openended recruitment period and lack of control regarding the allocation of subjects to groups based upon clinical interventions.

### Comparisons

In order to describe longitudinal microbiome progression free from obvious external modulators, a group of stable subjects was first identified from within the study population, who were minimally exposed to postnatal antibiotics. They (**Group 1**) were pragmatically defined as:-

- Having had no episodes of late-onset sepsis
- Had received only one course of antibiotics, in the immediate postpartum period
- Had >75% of stool samples obtained outside this period of antibiotic administration

This group would act as a baseline, against which microbiome development in other groups could be compared.

Two comparator groups were then identified from within the study population: 2a) *during* and 2b) *after* courses of antibiotics. Infants in Group 2a, *during* a period of antibiotic administration, were defined as:-

- The antibiotic course must be for an episode of LOS (*not* EOS)
- The antibiotic course must be preceded by at least five consecutive days without antibiotics
- Only one episode of antibiotic use was analysed per subject
- There must be at least three samples from within the period of antibiotic use
- Samples must be within the period 48 hours prior to commencing antibiotics (one pre-antibiotic sample only) and 48 hours following antibiotic cessation

From within Group 2a, a sub-group, 2b, was identified in whom microbiome development in the period *after* antibiotic cessation could be studied. This was defined as:-

- The period of assessment must occur in an antibiotic-free interval, following an episode of antibiotic use for LOS
- At least three samples from this antibiotic-free period must be available for analyses

Not all subjects in Group 2a could be studied as part of Group 2b, because there were too few samples in the period after antibiotics were stopped. No subjects in Groups 2a/2b were in Group 1, by virtue of antibiotic use for LOS. Subjects were allocated to their respective clinical groups following completion of recruitment, and prior to assessment of their microbiome composition.

Parameters of microbiome development were studied in these three groups, with comparison made between those receiving antibiotics for presumed LOS and those with no antibiotic exposure after the immediate post-partum period (*i*.*e*. Group 2a *vs*. Group 1; Group 2b *vs*. Group 1). Longitudinal diversity progression, and longitudinal progression of *Enterobacteriaceae* relative abundance were the two parameters which could be seen to follow roughly linear trajectories across the majority of subjects, and are quantitatively described. Other features of microbiome development in the three groups are noted, but were not appropriate for quantitative longitudinal analyses (due to grossly uneven distributions across subjects, or following inconsistent trajectories).

Preliminary analyses demonstrated that longitudinal diversity progression followed an approximately linear trajectory within three groups. Consequently, longitudinal diversity progression was described on an individual-level, as the rate of change of diversity with time. The mean/median of these individual diversity progressions was used as a summary measure to describe the progression within the wider group (*i*.*e*. Group 1/2a/2b).

Longitudinal development of *Enterobacteriaceae* relative abundance was similarly described. From an inspection of individual profiles, levels could be seen to either rise to a peak then fall, or fall from an initial peak. Consequently, for consistency, the rate of *Enterobacteriaceae* fall from its highest level was described on an individual level, as the percentage change in relative abundance with time. Again, the mean/median of these individual values was used to describe the wider groups.

Comparison was made between the summary measures of progression in Group 1 *vs*. Group 2a and Group 1 *vs*. Group 2b, using two-tailed weighted t-tests, or Mann-Whitney U-tests, as appropriate.

## Accessibility of sequencing data

The raw sequencing data and metadata were uploaded to a publicly accessible database (The NCBI Sequence Read Archive) under the BioProject accession code PRJNA605031.

## Supporting information

STROBE Checklist

## Data Availability

All data produced in the present study are available upon reasonable request to the authors.
The raw sequencing data and metadata were uploaded to a publicly accessible database (The NCBI Sequence Read Archive) under the BioProject accession code PRJNA605031.

## Acknowledgements

The work leading to this publication would not have been possible without the generous support of Barts Charity. Our additional thanks goes to the staff of the neonatal unit at Homerton University Hospital, for their hard work in the care of these babies and assistance in sample and data collection, and to the families and babies who participated, for their willingness to contribute to research, even at a difficult time.

## Notes

### Competing Interest Statement

The authors have declared no competing interest.

### Author Declarations

Ethical approval was granted by the London (Chelsea) Research Ethics Committee (Ref: 15/LO/1924).

## References

1. La Rosa PS, Warner BB, Zhou Y, et al. Patterned progression of bacterial populations in the premature infant gut. Proceedings of the National Academy of Sciences of the United States of America. 2014;111(34):12522–12527.

2. Butcher J, Unger S, Li J, et al. Independent of Birth Mode or Gestational Age, Very-Low-Birth-Weight Infants Fed Their Mothers’ Milk Rapidly Develop Personalized Microbiotas Low in Bifidobacterium. J Nutr. 2018;148(3):326–335.

3. Drell T, Lutsar I, Stsepetova J, et al. The development of gut microbiota in critically ill extremely low birth weight infants assessed with 16S rRNA gene based sequencing. Gut microbes. 2014;5(3):304–312.

4. Jacquot A, Neveu D, Aujoulat F, et al. Dynamics and clinical evolution of bacterial gut microflora in extremely premature patients. The Journal of pediatrics. 2011;158(3):390–396.

5. Cong X, Xu W, Janton S, et al. Gut microbiome developmental patterns in early life of preterm infants: Impacts of feeding and gender. PLoS ONE. 2016;11(4).

6. Arboleya S, Sánchez B, Solís G, et al. Impact of prematurity and perinatal antibiotics on the developing intestinal microbiota: A functional inference study. International Journal of Molecular Sciences. 2016;17(5).

7. Jia J, Xun P, Wang X, et al. Impact of Postnatal Antibiotics and Parenteral Nutrition on the Gut Microbiota in Preterm Infants During Early Life. Journal of Parenteral and Enteral Nutrition. 2019.

8. Blakey JL, Lubitz L, Barnes GL, Bishop RF, Campbell NT, Gillam GL. Development of gut colonisation in pre-term neonates. Journal of Medical Microbiology. 1982;15(4):519–529.

9. Zwittink R, Van Zoeren D, Martin R, et al. Development of the intestinal microbiota after short and long antibiotic treatment in latepreterm and term infants. Journal of Pediatric Gastroenterology and Nutrition. 2016;62:417.

10. Rooney AM, Timberlake K, Brown KA, et al. Each Additional Day of Antibiotics is Associated with Lower Gut Anaerobes in Neonatal Intensive Care Unit Patients. Clinical infectious diseases : an official publication of the Infectious Diseases Society of Americax. 2019.

11. Eastick K, Leeming JP, Bennett D, Millar MR. Reservoirs of coagulase negative staphylococci in preterm infants. Arch Dis Child Fetal Neonatal Ed. 1996;74(2):F99–104.

12. Hall MA, Cole CB, Smith SL, Fuller R, Rolles CJ. Factors influencing the presence of faecal lactobacilli in early infancy. Archives of disease in childhood. 1990;65(2):185–188.

13. Arboleya S, Sanchez B, Milani C, et al. Intestinal microbiota development in preterm neonates and effect of perinatal antibiotics. The Journal of pediatrics. 2015;166(3):538–544.

14. Greenwood C, Morrow AL, Lagomarcino AJ, et al. Early Empiric Antibiotic Use in Preterm Infants Is Associated with Lower Bacterial Diversity and Higher Relative Abundance of Enterobacter. The Journal of pediatrics. 2014;165(1):23–29.

15. Clark RH, Bloom BT, Spitzer AR, Gerstmann DR. Reported medication use in the neonatal intensive care unit: data from a large national data set. Pediatrics. 2006;117(6):1979–1987.

16. Hooper LV, Wong MH, Thelin A, Hansson L, Falk PG, Gordon JI. Molecular Analysis of Commensal Host-Microbial Relationships in the Intestine. Science. 2001;291(5505):881.

17. Stappenbeck TS, Hooper LV, Gordon JI. Developmental regulation of intestinal angiogenesis by indigenous microbes via Paneth cells. Proceedings of the National Academy of Sciences of the United States of America. 2002;99(24):15451–15455.

18. Kelly D, Campbell JI, King TP, et al. Commensal anaerobic gut bacteria attenuate inflammation by regulating nuclear-cytoplasmic shuttling of PPAR-gamma and RelA. Nature immunology. 2004;5(1):104–112.

19. Neish AS, Gewirtz AT, Zeng H, et al. Prokaryotic regulation of epithelial responses by inhibition of IkappaB-alpha ubiquitination. Science. 2000;289(5484):1560–1563.

20. Neal MD, Leaphart C, Levy R, et al. Enterocyte TLR4 mediates phagocytosis and translocation of bacteria across the intestinal barrier. Journal of immunology (Baltimore, Md : 1950). 2006;176(5):3070–3079.

21. Neal MD, Sodhi CP, Jia H, et al. Toll-like receptor 4 is expressed on intestinal stem cells and regulates their proliferation and apoptosis via the p53 up-regulated modulator of apoptosis. The Journal of biological chemistry. 2012;287(44):37296–37308.

22. Yazji I, Sodhi CP, Lee EK, et al. Endothelial TLR4 activation impairs intestinal microcirculatory perfusion in necrotizing enterocolitis via eNOS–NO–nitrite signaling. Proceedings of the National Academy of Sciences of the United States of America. 2013;110(23):9451–9456.

23. Kamdar S, Hutchinson R, Laing A, et al. Perinatal inflammation influences but does not arrest rapid immune development in preterm babies. Nature Communications. 2020;11(1):1284.

24. Bharucha T, Oeser C, Balloux F, et al. STROBE-metagenomics: a STROBE extension statement to guide the reporting of metagenomics studies. The Lancet Infectious Diseases. 2020;20(10):e251–e260.

25. Simpson EH. Measurement of Diversity. Nature. 1949;163:688.

26. Matthews JN, Altman DG, Campbell MJ, Royston P. Analysis of serial measurements in medical research. BMJ : British Medical Journal. 1990;300(6719):230–235.

27. Theil H. A rank-invariant method of linear and polynomial regression analysis. I, II, III Proceedings of the Royal Netherlands Academy of Sciences 1950;53:386-392, 521-525, 1397-1412.

28. Zimmermann P, Curtis N. The effect of antibiotics on the composition of the intestinal microbiota - a systematic review. The Journal of infection. 2019;79(6):471–489.

29. Fouhy F, Guinane CM, Hussey S, et al. High-throughput sequencing reveals the incomplete, short-term recovery of infant gut microbiota following parenteral antibiotic treatment with ampicillin and gentamicin. Antimicrob Agents Chemother. 2012;56(11):5811–5820.

30. Bhalodi AA, van Engelen TSR, Virk HS, Wiersinga WJ. Impact of antimicrobial therapy on the gut microbiome. The Journal of antimicrobial chemotherapy. 2019;74(Suppl 1):i6-i15.

31. Dardas M, Gill SR, Grier A, et al. The impact of postnatal antibiotics on the preterm intestinal microbiome. Pediatric research. 2014;76(2):150–158.

32. Bonnemaison E, Lanotte P, Cantagrel S, et al. Comparison of fecal flora following administration of two antibiotic protocols for suspected maternofetal infection. Biology of the neonate. 2003;84(4):304–310.

33. Cotten CM, Taylor S, Stoll B, et al. Prolonged Duration of Initial Empirical Antibiotic Treatment Is Associated With Increased Rates of Necrotizing Enterocolitis and Death for Extremely Low Birth Weight Infants. Pediatrics. 2009;123(1):58.

34. Kuppala VS, Meinzen-Derr J, Morrow AL, Schibler KR. Prolonged initial empirical antibiotic treatment is associated with adverse outcomes in premature infants. The Journal of pediatrics. 2011;159(5):720–725.

35. Abdulkadir B, Nelson A, Skeath T, et al. Stool bacterial load in preterm infants with necrotising enterocolitis. Early Human Development. 2016;95:1–2.

36. Torrazza RM, Ukhanova M, Wang X, et al. Intestinal microbial ecology and environmental factors affecting necrotizing enterocolitis. PLoS One. 2013;8(12):e83304.

37. Mai V, Young CM, Ukhanova M, et al. Fecal microbiota in premature infants prior to necrotizing enterocolitis. PloS one. 2011;6(6):e20647.

38. Gribar SC, Sodhi CP, Richardson WM, et al. Reciprocal expression and signaling of TLR4 and TLR9 in the pathogenesis and treatment of necrotizing enterocolitis. Journal of immunology (Baltimore, Md : 1950). 2009;182(1):636–646.

39. Bry L, Falk PG, Midtvedt T, Gordon JI. A Model of Host-Microbial Interactions in an Open Mammalian Ecosystem. Science. 1996;273(5280):1380.

40. Hooper LV, Stappenbeck TS, Hong CV, Gordon JI. Angiogenins: a new class of microbicidal proteins involved in innate immunity. Nature immunology. 2003;4(3):269–273.

41. Costeloe K, Bowler U, Brocklehurst P, et al. A randomised controlled trial of the probiotic Bifidobacterium breve BBG-001 in preterm babies to prevent sepsis, necrotising enterocolitis and death: the Probiotics in Preterm infantS (PiPS) trial. Health Technol Assess. 2016;20(66):1–194.

42. Tobias J, Olyaei A, Laraway B, et al. Bifidobacterium infantis EVC001 Administration Is Associated With a Significant Reduction In Incidence of Necrotizing Enterocolitis In Very Low Birth Weight Infants. The Journal of pediatrics. 2022.

43. Gibson MK, Wang B, Ahmadi S, et al. Developmental dynamics of the preterm infant gut microbiota and antibiotic resistome. Nature Microbiology. 2016;1(4):16024.

44. Raveh-Sadka T, Firek B, Sharon I, et al. Evidence for persistent and shared bacterial strains against a background of largely unique gut colonization in hospitalized premature infants. ISME Journal. 2016;10(12):2817–2830.

45. Leggett RM, Alcon-Giner C, Heavens D, et al. Rapid MinION profiling of preterm microbiota and antimicrobial-resistant pathogens. Nature Microbiology. 2020;5(3):430–442.

46. Ariefdjohan M, Savaiano D, Nakatsu C. Comparison of DNA extraction kits for PCR-DGGE analysis of human intestinal microbial communities from fecal specimens. Nutrition journal. 2010;9:23.

47. Nocker A, Sossa-Fernandez P, Burr MD, Camper AK. Use of Propidium Monoazide for Live/Dead Distinction in Microbial Ecology. Applied and Environmental Microbiology. 2007;73(16):5111–5117.

48. Young GR, Smith DL, Embleton ND, et al. Reducing Viability Bias in Analysis of Gut Microbiota in Preterm Infants at Risk of NEC and Sepsis. Frontiers in Cellular & Infection Microbiology. 7:237.

49. R: A language and environment for statistical computing [computer program]. R Foundation for Statistical Computing 2019.

50. Callahan BJ, McMurdie PJ, Rosen MJ, Han AW, Johnson AJA, Holmes SP. DADA2: High-resolution sample inference from Illumina amplicon data. Nature Methods. 2016;13:581.

51. Wong K, Fleming P, Millar M, et al. The Construction of the Curated Preterm Gut Microbiome Database (PGMD). FEMS; July 9-13th 2017, 2017; Valencia.

52. Davis NM, Proctor DM, Holmes SP, Relman DA, Callahan BJ. Simple statistical identification and removal of contaminant sequences in marker-gene and metagenomics data. Microbiome. 2018;6.

